# Assessing the intrinsic and extrinsic drivers and targeting the observed resilience of malaria in northwestern and southern Tanzania: A protocol for a cross-sectional exploratory study

**DOI:** 10.1101/2020.05.05.20091330

**Authors:** Mercy G. Chiduo, Celine I. Mandara, Susan F. Rumisha, Frank Chaky, Filbert T. Francis, Bruno P. Mmbando, Misago D. Seth, Daniel P. Challe, Leah Ndekuka, Isolide S. Massawe, Edwin Liheluka, Williams H. Makunde, Athanas D. Mhina, Vito Baraka, Method D. Segeja, Yahya A. Derua, Berndard M. Batengana, Paul M. Hayuma, Gineson Nkya, Rashid A. Madebe, Masunga C. Malimi, Renata Mandike, Sigsbert Mkude, Fabrizio Molteni, Ritha Njau, Ally Mohamed, Deus S. Ishengoma

## Abstract

**Background:** Despite high coverage and successes in malaria control strategies, some areas of Tanzania have indicated stagnantion or revesal of malaria burden. In malaria research, most studies are designed to assess drivers of malaria transmission focusing only on one dimension, single location while very few studies assess multiple components and their interactions at once. This article describes the protocol used to assess intrinsic and extrinsic drivers of persistent malaria transmission (hotsposts) in four regions from northwestern (Geita and Kigoma) and southern (Ruvuma and Mtwara) Tanzania.

**Methods:** This cross-sectional study was conducted between July and November 2017 in eight districts (two from each region). Based on the health facilities records, two villages were selected from each district. The study assessed five components individually and their linkages: socio-economic and malaria risk factors, parasitological, entomological, socio-anthropological and health system factors. Households (HHs) and household members were enumerated, socio-economic status and risk factors associated with malaria transmission were assessed. A total of 120 HHs were sampled from each village where malaria testing using rapid test and microscopy were done and blood spots on filter papers for genetic studies were collected. Heads of HHs were interviewed to capture information on knowledge, attitude, practice and beliefs towards malaria and its control. Assessment of adult mosquitoes in 25 HHs in each village and complimented with assessment of immature mosquitoes through larvae was conducted. The performance of the health system was assessed with respect to Information on availability, accessibility, affordability and quality of malaria prevention and case management services were collected from these health facilities.

**Discussion:** The proposed analysis plan and results from this study are expected to determine factors potentially responsible for persistence of malaria (hotspots) in the study areas. Rather than the traditional methodology of focusing on one metric, the approach proposed here triangulates observations from all five components, highlighting understanding of potential drivers while studying their complex interactions and map spatial heterogeneity. This study will provide an important framework and data which will guide future studies and malaria surveillance in Tanzania and other malaria endemic countries.

## BACKGROUND

Recent reports have shown a dramatic decline in malaria around the world and across sub-Saharan Africa (SSA)(1). However, the 2015-2016 Tanzania demographic and health survey and malaria indicator survey (TDHS-MIS) showed re-emergence of malaria with an increase from its 2012 prevelence (among children 6-59 months old) of 9.8%(2) to 14.8%(3). This highly heterogenous disease continues to place a heavy burden in most parts of mainland Tanzania, including the southern and north western regions with prevalence rates in some regions exceeding 50% and high mortality rates in young children(3,4). Although the results from the different national surveys have demonstrated an upward trend in intervention coverage, particularly long-lasting insecticidal nets (LLINs) (>80%) and a substantial increase in use rate (66%), this is not reflected in the trends of disease burden in some parts(3). It is evident that the current interventions pioneered by the Tanzanian national malaria control programme (NMCP) might be insufficient in some areas and less likely to lead to the elimination target of malaria in these poverty stricken areas. In the fight against malaria and in order to reduce the disease burden, there might be a need for i) complimentary strategies; and ii) strategic and innovative deployment of available tools, in order to reach the national and international malaria control/elimination goals.

Recent successes in malaria control efforts relying on insecticide-based vector control interventions such as LLINs (5–11) and indoor residual spraying (IRS) (12–15), are threatened by rapid expansion of insecticide resistance in vector population. This has affected many countries in Africa, and is mounting pressure on the advancing global malaria elimination efforts(16–20). To make the long term goal of malaria elimination in Africa a reality, it is important that the effectiveness of existing control interventions is maintained and supplemented with additional proven strategies for vector control, case management and behavioural change. Larval control and house improvement could be important interventions in both urban and rural settings, particularly in areas where majority of the population are at risk(21–25). In addition, improved longitudinal quality assured disease/vector surveillance systems, such as the community-based trapping system have become increasingly necessary for identifying and mapping the densities of sparse residual vector populations and for monitoring and evaluating the impact of interventions upon them (26–31).

The intensity of malaria transmission and the disease burden are influenced by a complex array of environmental, ecological, socio-demographic and climate factors and this complex interplay of biotic and abiotic factors should be taken into account in an attempt to assess the impact of the recent malaria interventions. Traditional approaches, assessment of factors related to transmission and burden is usually designed focusing on few components without considering the interlinks and interactions, which play between such attributes. This is mainly due to financial or logistic constraints, but also partly to ignoring or failure to fully consider the importance of the existing interlinks among different factors responsible for and maintenance of malaria transmission. Congruent temporal and spatial data over long periods are critical for determining key factors affecting malaria transmission at micro and macro-geographical levels, but such long-term data is commonly lacking. This results into limited evidence with high level of uncertainty which sometimes fail to provide an accurate measure of the magnitude of the disease risk and guidance on stratification of disease burden which is a prerequisite in setting up control strategies with most impact.

Previous surveys have shown variation in malaria prevalence in areas with small proximities and factors related to such patterns are not clearly known(3,32,33). In the absence of longitudinal data, stratification of malaria transmission can alternatively be attained through macro and micro-mapping of malaria transmission and disease burden, and documenting key factors responsible for heterogeneity of malaria. That need to cover important aspects including community-based indicators, profile of the disease epidemiology, coverage and utilization of interventions, vector biology and behaviours, and performance and readiness of the exisiting health system in disease management.

This article describes the protocol used to determine the intrinsic and extrinsic drivers of persistent malaria burden in selected areas of Tanzania mainland. It aimed at providing data on different factors which could be potentially driving the current pattern of malaria transmission and are possibly associated with persistent and/or reduced malaria burden and the heterogeneity currently observed in Tanzania. The findings will be used to support NMCP and its stakeholders in the stratification of malaria burden and developing targeted control interventions required to push malaria further down to reach elimination target by 2030. The data will also support future studies/surveillance of persistent malaria burden in different parts of Tanzania.

### Study objectives

The main objective of this assessment was to determine the intrinsic and extrinsic drivers of persistent malaria burden in selected parts of Tanzania. Specifically, focusing on testing existence of micro scale spatial variations of parasite prevalence, the distribution, type and abundance of malaria vectors. Addition to that, the study assessed socio-economic, anthropological and health system factors associated with persistence of malaria in regions which had high level of interventions in the past decade. Interlinks between the five components will be studied to understand their interactions and draw conclusions which have to be taken into account when assessing these relationships.

## Methods

### Study Sites

This study covered eight districts from four regions; Geita, Kigoma, Mtwara and Ruvuma regions (Figure 1), with persistently high malaria transmission as shown by different surveys conducted in Tanzania between 2011 and 2017. Such surveys include the Tanzania HIV and Malaria indicator survey (THMIS 2011-12) (32), Tanzania demographic and health survey – Malaria Indicator survey (TDHS-MIS 2015-2016) (34) and the 2015/2016 school malaria prevalence surveys (SMPS) (35) (Figures 2 and 3).

**Figure 1:**
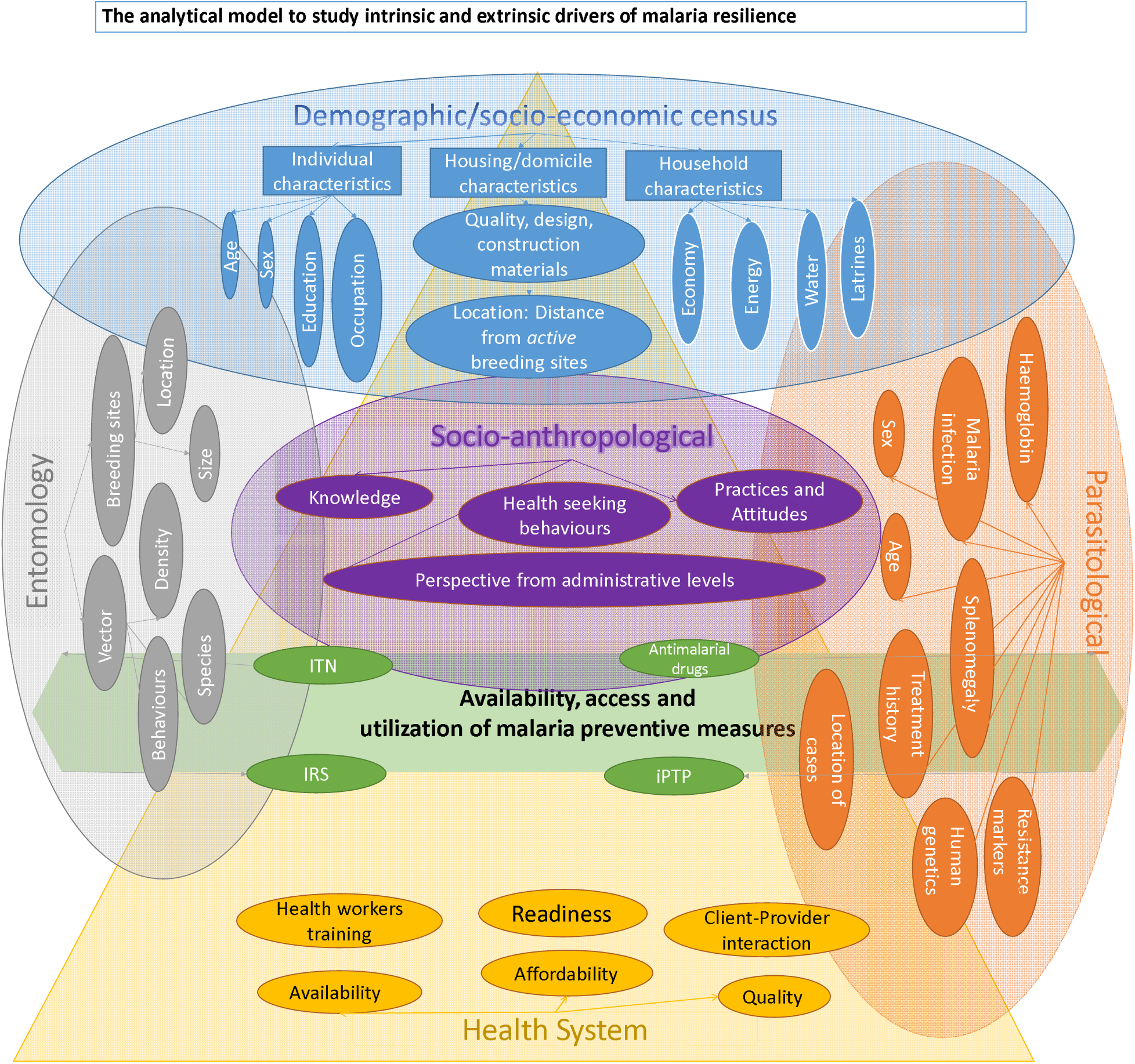
Map of Tanzania showing the location of the study regions, districts and villages

**Figure 2:**
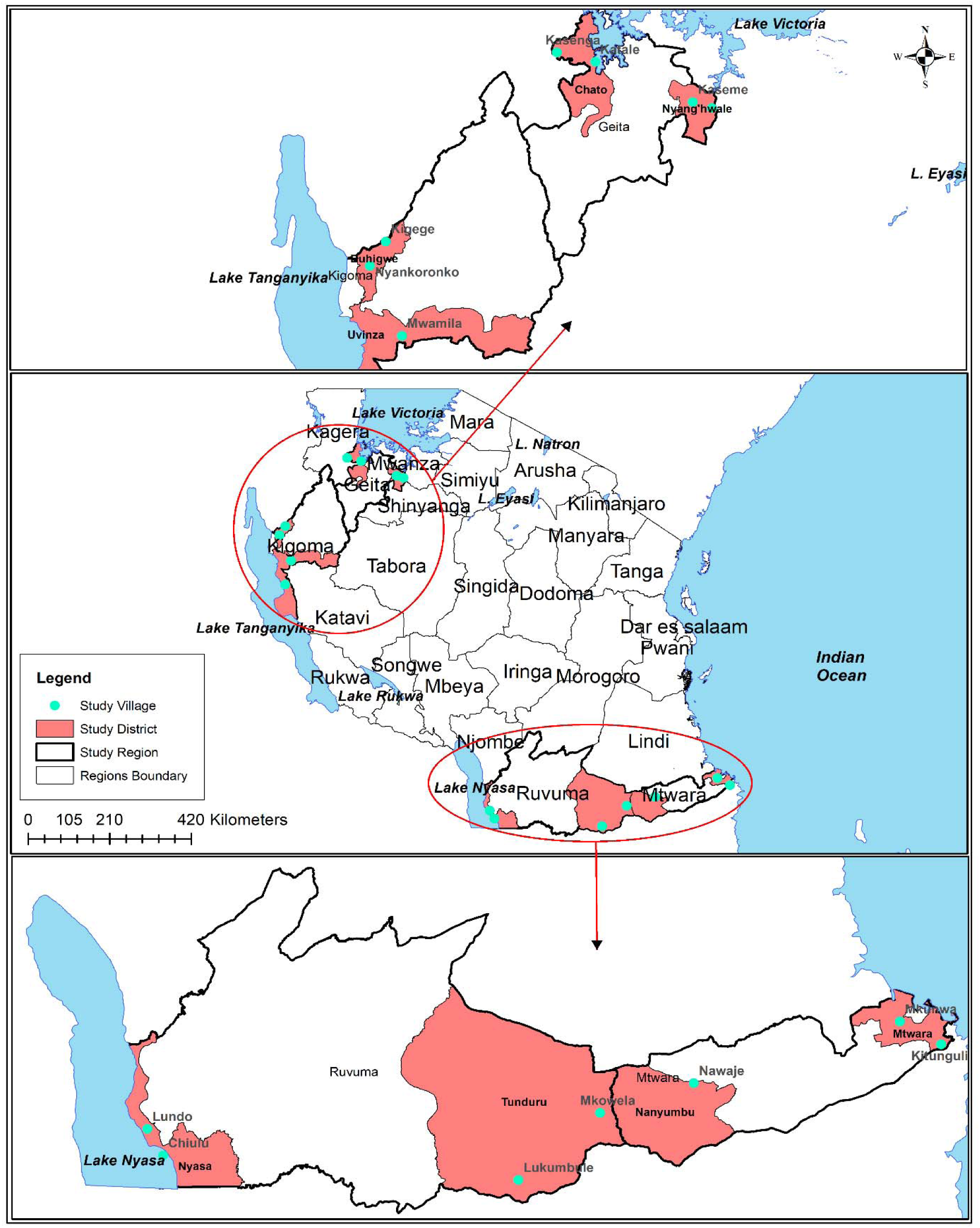
Maps of Tanzania showing malaria prevalence in children aged 0-59 months by region Source: MIS 2007/08, 20011/12 and 2015/16 (2,3,36)

**Figure 3:**
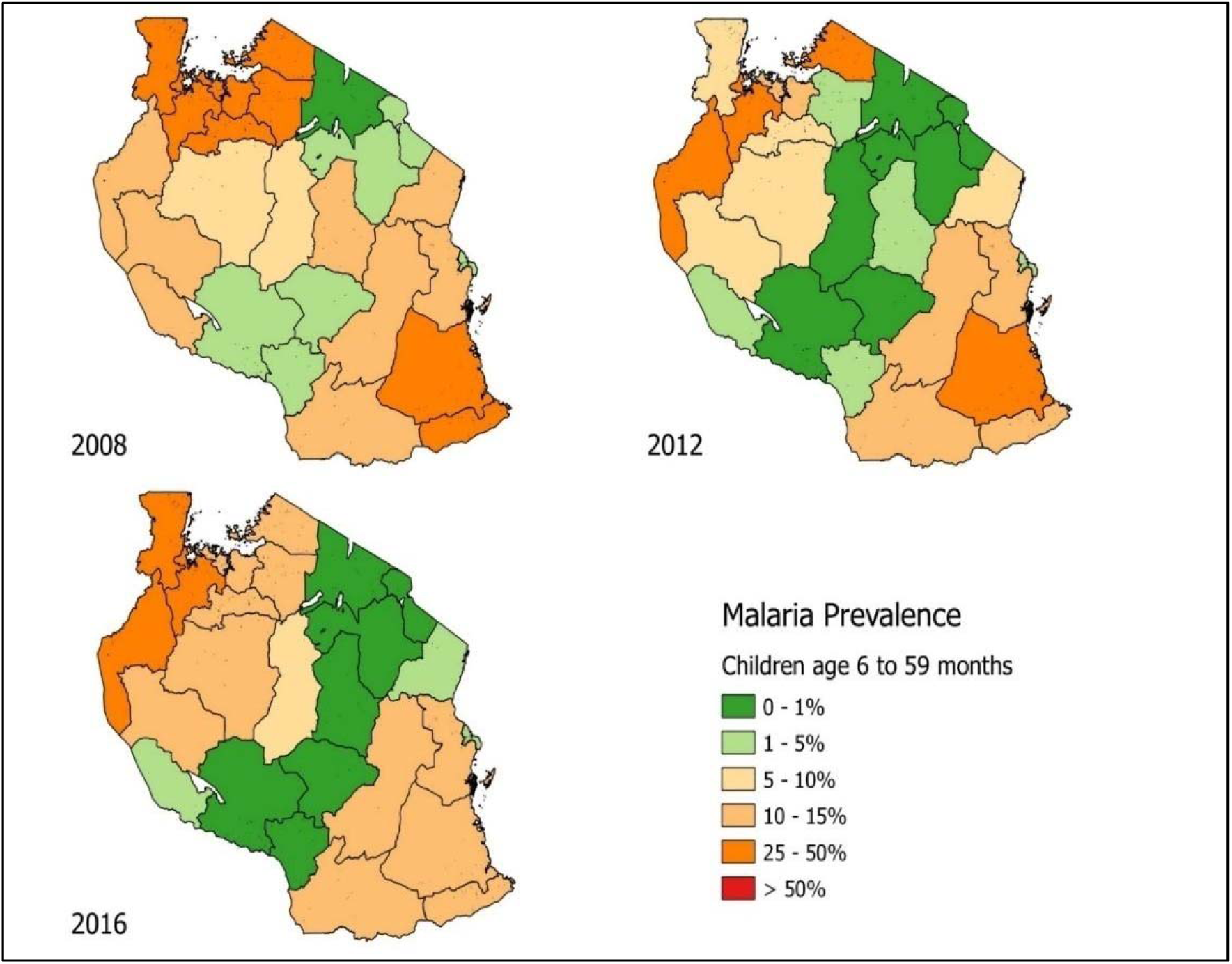
Map of Tanzania showing malaria prevalence among school children based on the SMPS of 2015/16 (35)

Based on the 2011-12 THMIS, the highest malaria prevalence (by malaria Rapid Diagnostic Test(mRDT)) was observed in the Southern (20.6%), Western (16.7%) and Lake (14.8%) zones(2). A similar pattern was observed 3 years later during the 2015-16 TDHS-MIS which showed that the highest prevalence was within the same zones; Western (27.7%), Lake (23.5%) and Southern (18.8%)(3). Regions with high (>10%) prevalence during the 2011-12 survey included (in descending order); Geita (31.8%), Lindi (26.3%), Kigoma (26.0%), Mara (25.4%), Mwanza (18.6%), Mtwara (17.4%), Morogoro (13.0%) and Ruvuma (12.0%)(2). The same set of regions presented high prevalence in 2015-16 in additional to Kagera, Pwani, Tabora, Shinyanga, Mwanza, Katavi and Simiyu(3). The results of SMPS brought a similar set of zones and regions affected mostly with malaria. Regions with the highest prevalence included Geita (53.7%), Pwani (48.4%), Mwanza (40.0%), Katavi (39.9%), Mara (36.4%), Mtwara (36.2%), Shinyanga (35.1%), Kagera (31.1%), Simiyu (31.0%), Lindi (30.3%), Kigoma (30.3%), Tabora (30.0%) and Ruvuma (22.6%)(35). The findings of these surveys indicated that some regions consistently maintained high malaria transmission in the past 5 years, including Geita (Lake zone), Ruvuma and Mtwara (Southern zone) and Kigoma (Western zone) (Figure 1).

Thus, four regions were selected for the study and two districts with high and low malaria prevalence based on the SMPS of 2015/2016 were selected from each region (Figure 1); Buhigwe and Uvinza (Kigoma), Mtwara DC and Nanyumbu (Mtwara), Nyasa and Tunduru (Ruvuma) and Nyang’hwale and Chato (Geita). Districts in Geita region were chosen with an added criterion of taking part in IRS programme which was done shortly before this study. Two villages with high malaria burden, based on the number of cases reported by health facilities in 2016, were selected from each district, making 16 villages. Health facilities serving the respective villages were also visited for the health system survey.

### Study design

This was a cross-sectional survey which evaluated entomological, parasitological, health systems, socio-economic and socio-anthropological components (Figure 4).

**Figure 4.**
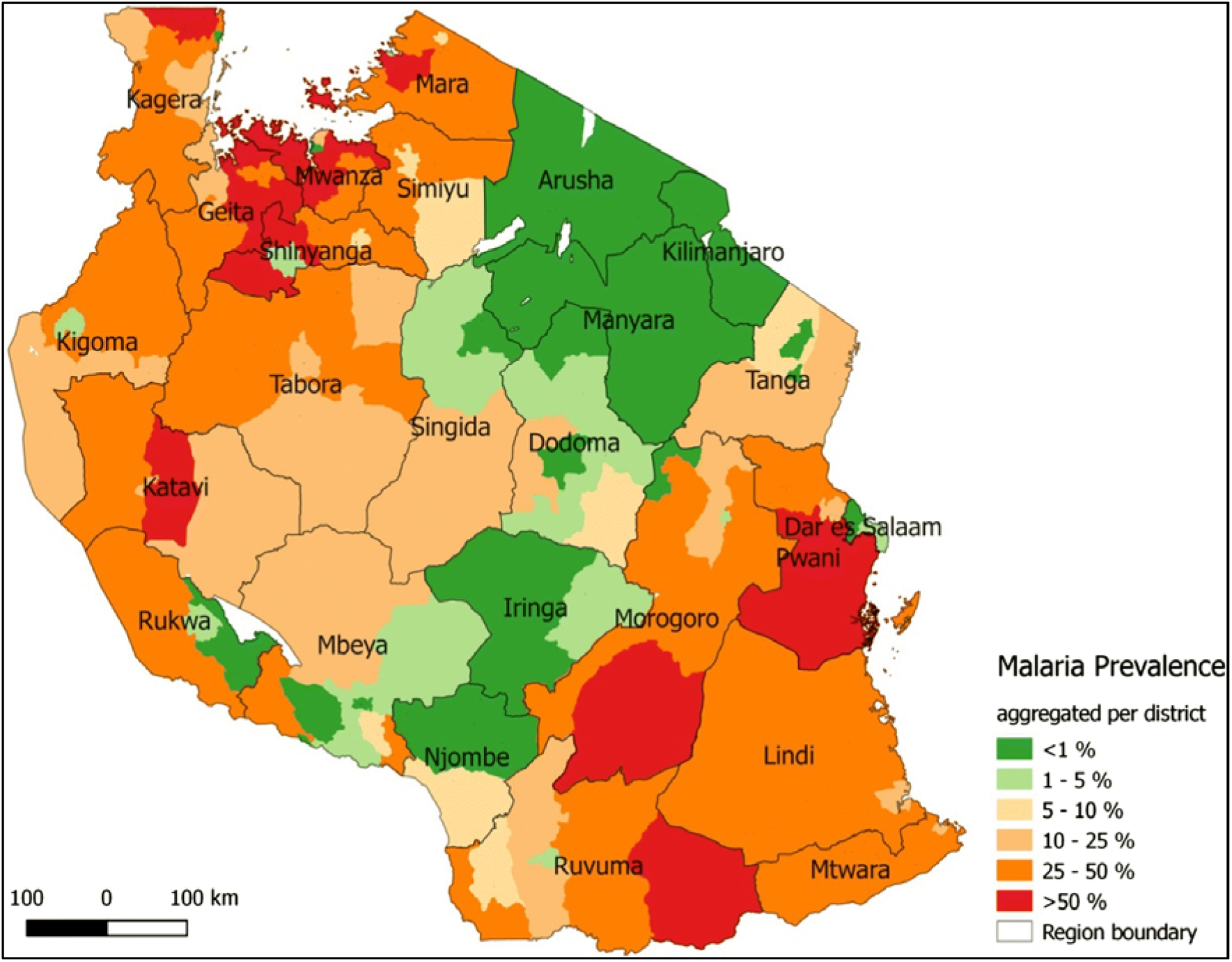
Flow chart showing the study design.

### Plannning and organization of the study teams

Training of the teams on the study protocol and the tools was done before initiation of the study; and was done separately according to the component to be implemented. Dry runs were conducted in Magoda village in Muheza district, Tanga region. Community members of Magoda village have experience of taking part in different surveys of malaria transmission for the past three decades (37). The demographic and socio-economic (census) team conducted first the dry run, other teams conducted the dry runs one week later using the data generated by the census team. The exercise was followed-up by a post-test assessment to sort out any issues encountered during the dry runs before data collection was launched. The main study was conducted between July and November 2017 following a similar order. In each village, the teams conducted the survey in 3 to 4 days and the entire survey took about 65 days.

### Sample size, sampling procedures, and collection of data and samples

To create a sampling frame, a demographic census was undertaken and all households (HHs) from the selected villages were enumerated, and each member of the HH was given a unique identification number. Systematic sampling was used to identify HHs to be included in other components of the study as described below.

### Demographic and socio-economic survey

A demographic census was undertaken in the selected villages whereby all HHs (between 200 to 700HHS per village) were enumerated and assessed for socio-economic status (SES) and risk factors associated with malaria transmission. Demographic data including name, sex, date of birth, education status and occupation were collected for all members residing in each HH. Malaria risk mapping was undertaken, and it included collection of data on the housing quality based on construction materials, presence of eaves and open windows, location of the HH in the village and environmental features associated with malaria transmission (e.g. mosquito breeding sites). Furthermore, information on malaria prevention strategies [e.g. screened windows, availability and use of bed nets (ITNs and LLINs), IRS and intermittent presumptive treatment in pregnancy using SP (IPTp-SP)] were collected. The SES of each HH was also assessed and it focused on ownership of assets such as livestock, farming land, electronic equipment (e.g. mobile phones, radio, television), means of transport (e.g. bicycle, motorcycle), source of energy for cooking and lighting, source of drinking water and type of latrines.

### Parasitological survey

A total of 120 HHs from each village were sampled from the census list and all members from the selected HHs were enrolled in the parasitological survey that was conducted at a central point. A comprehensive parasitological assessment was conducted to obtain baseline data on malaria transmission intensity in the study population. A parasitological questionnaire was used to collect information on demographics, malaria prevention, clinical and treatment history. Additionally, vital signs and anthropometrics measures were taken, splenomegaly was evaluated for children up to 12-years. Blood samples were collected by finger prick, thin and thick smears were prepared, malaria rapid test (mRDT) (Carestart™ Somerset, NJ USA) was done for detection of malaria parasites and haemoglobin levels were evaluated using HaemoCue® machines. Dried blood spots (DBS) on filter papers were collected for further analysis of malaria parasites and host genetic factors that might affect susceptibility/resistance to malaria parasite infection and clinical disease. Participants with mRDT positive results were treated according to the National malaria treatment guidelines(38) and participants thought to need further investigation and management were referred to the nearest health facility.

### Socio-anthropological survey

A structured questionnaire was used to collect quantitative data while qualitative data was collected through focus group discussions (FGDs) and in-depth interviews (IDIs) using interview topic guides. Heads of the HHs which were involved in the parasitological survey were interviewed to capture information on knowledge, attitude, practices and beliefs (KAP&B) towards malaria and its control. Specifically, respondents provided information on key issues such as malaria transmission, signs, symptoms and treatment; malaria prevention (e.g. use of bed-nets and chemoprevention such as IPTp) and health seeking behaviour. For the qualitative study, two FGDs were conducted in each village, whereby each FGD included 8-12 individuals aged >18 years (male and female participants separately). The information collected during FGDs included; knowledge on malaria (its transmission, signs, symptoms and treatment), community’s practices and attitudes on malaria prevention, e.g. use of bed-nets, use of antimalarial drugs and health seeking behaviour. Participants of the FGDs were randomly selected from HHs which were not involved in the parasitological survey and KAP&B interviews. Indepth interviews were also undertaken with at least two officials involved in malaria control at the district level, including the district medical officer and/or district-malaria coordinators/focal persons. Repondents provided information on availability, accessibility and utilization of malaria prevention, treatment and control services in their areas.

### Entomological survey

A sub-sample of 25 HHs were selected randomly (from the census list) in each of the study villages for entomological assessments. The selected HHs had open eves, were close to the breeding sites and with medium size windows and/or with no mosquito netting material. Among the 25 HHs; 20, 10 and 5 HHs were randomly selected for collection of indoor host seeking malaria vectors using the Centres for Disease Control (CDC) light traps (John W Hock Co, Gainesville, FL, USA), indoor resting malaria vectors using pyrethrum spray catch and outdoor biting malaria vectors using tent traps, respectively. Assessment of immature mosquitoes was also done through larvae search. Details of each collection method used are provided below:

#### Light Trapping

After obtaining a written informed consent from the home owner, a CDC light trap was installed at the foot of a sleeping place occupied by a family member sleeping under an insecticide-treated bed-net as described previously (39). In brief, trapping was done in 20 HHs from each village for three consecutive nights and the traps were set between 18.00 and 19.00 hours, and retrieved the following morning between 06:00 and 07:00 hours.

#### Pyrethrum Spray Catches

Spray catch collection was done for one day in 10 HHs from each study village. The HHs used here were a sub-sample of the same HHs used for CDC light trapping. Mosquitoes were collected by spraying a non-residual crude pyrethrum solution premixed with kerosene at the concentration of 0.5%. Food and small items were removed from the houses and white cotton sheets were spread to cover the floor and large furniture. Ten to fifteen minutes after spraying, researchers collected knocked down mosquitoes and preserved them in petri-dishes for later identification and score of their physiological status.

#### Furvela Tent Trapping

These were used for three nights consecutively in 5 HHs from each village. Furvela tent traps consisted of a tent and a CDC light trap with the light bulb removed. A volunteer slept in the tent with a small opening in the tent door to allow host odours to escape and attract mosquitoes as previously described (40). The traps were set each night between 18:00 and 19:00 hours and retrieved the following morning between 06:00 and 07:00 hours.

#### Larval Surveillance

Mosquito larval breeding habitats were mapped using global positioning system (GPS) in Open data kit (ODK) (GPS; Trimble Geoexplorer II, USA) while Larvae search was conducted using the World Health Organization (WHO) 350 ml standard mosquito dipper. Depending on the size of the aquatic habitat, five to twenty dips were taken from each larval habitat after physically inspecting the habitat for the presence or absence of mosquito larvae. Five dips were taken in small larval habitats of ≤1 m^2^, 10 dips for medium-sized habitats (2–15 m^2^) and 20 dips for relatively large habitats (> 15 m^2^). The immature mosquitoes were classified into three categories as; early instars (first and second stage larvae), late instars (third and fourth stage larvae) and pupae, after which they were sorted by genus, counted and recorded.

#### Mosquito identification

Mosquitoes collected were transported to the field laboratory which was set up in each village (within a school or village administration office) and identified to species using morphological criteria (41). Malaria vectors were further classified according to their abdominal status as unfed, blood-fed, semi-gravid or gravid. Female mosquitoes (*Anopheles gambiae* complex and *An. funestus* group) were stored in individual Eppendorf tubes containing silica gel desiccant for sibling species identification, detection of sporozoite and molecular markers of insecticide resistance. All non-malaria mosquitoes were identified, counted and discarded.

### Health system survey

The health system (HS) assessment survey targeted all health facilities serving the study villages including Accredited Drug Dispensing Outlets (ADDO), dispensaries, health centres and district hospitals where village members were getting initial consultations and/or refferal services. The facilities include those located within the study villages or nearby. The assessment was based on the WHO recommended guidelines on service provision assessment for malaria (SPAM)(42) which collects information on availability, accessibility, affordability and quality of malaria prevention and case management services. Data collection tools, adopted from the WHO guidelines, were used for this assessment. These included, Health facility Profile, Consistency check, Antenatal clinic (ANC), Out-patient department (OPD), Inpatient department (IPD), Laboratory and Pharmacy questionnaires. Data were collected from the facilities through observations and documentary review, health workers interview and exit interviews with clients. A critical assessment of different units/sections and departments at the health facilities (including outpatient and inpatient departments, laboratory and reproductive child health – RCH units) and within ADDOs was done. Health workers interviewed included the facility in-charges and heads of units while exit interviews were held with patients’/care-givers who were attended at the mentioned units on the day of the visit (interviewees were selected based on history of fever or diagnosis of malaria). In summary, observations were done for 12 patients attended at each facility including; 5 malaria patients from outpatient department, 5 from RCH and 2 patients who received artesunate injection for treatment of severe/complicated malaria.

## Laboratory Analyses

### Detection of malaria parasites by microscopy and Polymerase chain reaction (PCR)

Blood smears and DBS sample were sent to laboratory in Tanga for further analysis. The blood slides were dried at room temperature, thin film fixed with 99% Methanol and the smears were stained with 5% Giemsa solution for 45 minutes the following day while in the field. Stained blood slides were then packed and shipped to the laboratory in Tanga where the smears were examined under high power objective (oil immersion objective) for identification and quantification of malaria parasites. Asexual and sexual parasites were counted against 200 and 500 white blood cells (WBCs), respectively. Parasite density was obtained by multiplying the counts by 40 or 16 for asexual and sexual parasites, respectively; assuming each micro litre of blood contained 8000 WBCs(43). A smear was declared negative after examining 200 high power fields.

DNA was extracted from all microscopy positive DBS samples using QIAamp DNA Mini Kits (QIAGEN, Valencia, CA, USA) as described by the manufacturer, and stored at −20°C before analysis. Other samples will be extracted later using the same methods and the photo-induced electron transfer PCR (PET-PCR) assay will be used to detect the presence of sub-microscopic infections. The assay relies on self-quenching primers for the detection of *Plasmodium spp* using *Plasmodium* genus and species specific primers as previously described (44).

### Analysis of malaria vectors

Preserved mosquito samples were shipped to the laboratory in Tanga for further processing and analysis. Polymerase chain reaction for identification of sibling species of the An. *gambiae* complex and *An. funestus* group was conducted on a sub-sample of adult mosquito selected randomly from each village. Genomic Deoxyribonecleic acid (gDNA) was extracted from wings or legs of a female *An. gambiae* complex and An. *funestus* group according to methods previously described (45). The DNA samples were used for identification of sibling species of An. *gambiae* complex and An. *funestus* group using species specific diagnostic primers for An. *gambiae* complex (*An. gambiae ss., An. arabiensis, An. quadriannulatus, An. melas, An*. merus) (46). For the sibling species of the An. *funestus* group, the samples were identified based on species-specific primers in the internal transcribed spacer 2 (ITS2) region of the recombinant DNA (rDNA) genes, using a method previously developed to identify An. *funestus, An. vaneedeni, An. rivulorum, An. leesoni*, and An. *parensis* (47). Genotyping results were analysed using MXPro software (Agilent technologies, Santa Clara, CA, USA). Further analysis of malaria vector will be performed later including i) use of real time PCR (using TaqMan assays) to detect mutations in the knock down resistance (*kdr) gene* associated with pyrethroid resistance and determine the presence of *kdr* east and west genotypes in An. *gambiae* complex (48); ii) single nucleotide polymorphisms (SNPs) genotyping to assess the population structure of mosquitoes and additional markers of insecticide resistance; and, iii) Detection of *P. falciparum* circumsporozoite protein (Pf-CSP) and source of blood meal (for blood fed vectors) on a sub-sample of mosquitoes (An. *gambiae* complex and An. *funestus* group) by enzyme-linked immunosorbent assay (ELISA) technique as previously described(49).

### Analysis of parasite population structure and molecular markers of antimalarial resistance

The samples were genotyped for parasite population structure at the University of North Carolina (USA) using molecular inversion probes (MIP) protocol as described earlier (51). The data has been used to assess the population structure and other population genetics metrics in the parasite populations from the study areas (Mosers et al. Manuscript in Preparation). The assay based on MIPs was also used to detect molecular markers of drug resistance. The analysis will focus on markers of artemisinin resistance (P. *falciparum* kelch 13, k-13), partner drugs particularly lumefantrine and amodiaquine (P. *falciparum* multidrug resistance – *Pfmdn* gene) while piperaquine (*Plasmepsin 2 and* 3) will be assessed later. Markers of resistance to the old drugs which were previously used in Tanzania – chloroquine (chloroquine transporter gene *Pfcrt*) and sulphadoxine/pyrimethamine – SP (dihypteorate synthase and dihydrofolate reductase, *Pfdhps* and *Pfdhfr* genes) were also assessed. The *Pfmdn* copy number variation will be detected by real-time PCR following a previously described assay(52).

Additional analysis was done to detect the levels of *P. falciparum* Histidine rich protein 2 (HRP2) and the prevalence of *hrp2/3* gene deletion and their impacts on the performance of HRP2based mRDTs in Tanzania. Analysis of HRP2 will be done using a bead luminex assay described earlier (Rogier et al)(53), while the gene deletion will be assessed using the molecular assay that has been optimized at CDC (Bakari et al. Manuscript in Preparation). Polymorphisms in the *hrp2/3* genes was analysed by Sanger sequencing as previously described (53).

### Human genetic analyses

Human DNA will be extracted from DBS and analysed for polymorphisms in different genes which are known to be associated with susceptibility/ resistance to malaria. Such analyses will target sickle cell (β-globin), a-thalasemia (α-globin), glucose 6-phosphate dehydrogenase (G6PD) genes and others polymorphisms which will be deemed relevant.

### Data management

Quantitative data was collected using tablets which were connected through internet to the central server located at NIMR Tanga Centre. The ODK software was used to create the database and data collection applications. The data manager reviewed the data on a daily basis for precision and consistency. Queries generated on a daily basis were sent back to the head of each study team for resolution. Data cleaning and validation was done continuously and daily/weekly as well as periodic reports generated. Final cleaning was done after the survey and analysis is being performed according to the data analysis plan which was prepared at the beginning and finalized at the end of the study.

Qualitative data were collected using tape recorders and later the audio files were stored in the computer. Audio files were transcribed from Kiswahili and translated into English language. Thematic approach was used for data analysis.

### Data analysis

The Analytical framework and modelling approaches which will be used for the study is shown in figure 5.

**Figure 5.**
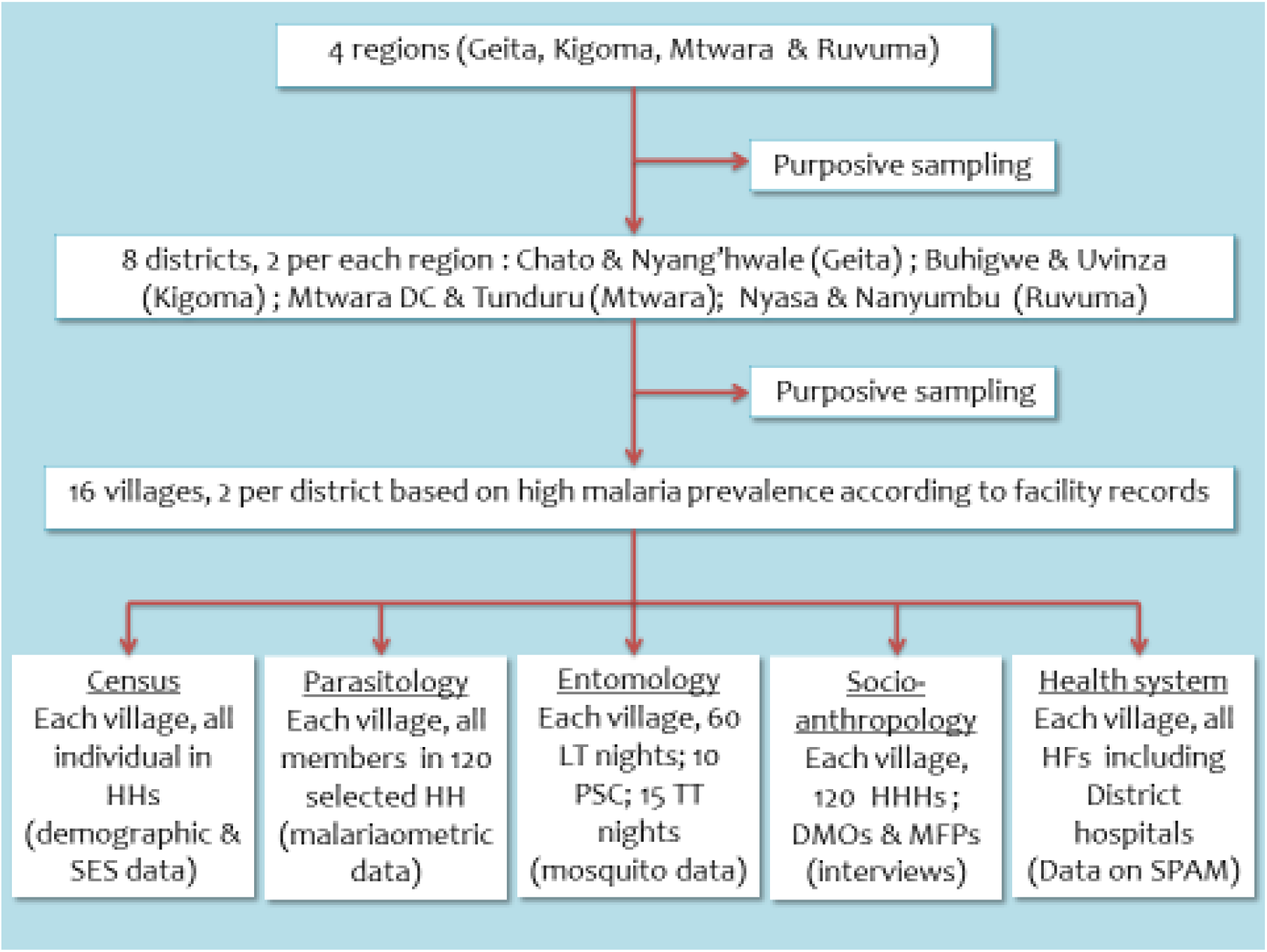
The Analytical framework and modelling approaches

The proposed analytical framework for determining the intrinsic and extrinsic drivers of malaria resilience can be derived starting from any of the five components depending on what action need to be taken or from which point of view implementation of the action targets. The model in Figure 5 illustrates the interlinks that exists between these components. Health system is assumed to affect all components, but with different magnitude and assumed to impacts more socio-anthropological and the parasitological factors. Availability, access and utilization of malaria preventive measures is a across-cutting component between HS, entomology and parasitology, and slightly between anthropological aspects, with more weight given to its linkage with performance of the HS and affects the entomological and parasitological components.

Prior to analysis, the following will be done: i) potential indicators/measures to be derived from each component will be identified. A list with details of data to be used in calculation of the indicators and what information is expected to contribute to the analytical framework will be created; ii) information from demographic/socio-economic census, available for all individuals in all study villages, will be used to create a community-based vulnerability index to malaria resilience in the study districts. The index will be build based on “who are we dealing with” -concept, i.e. the study population. A sensitivity analysis will be done to assess how the index change when adding information from other components. Malaria prevalence levels (at finest scale possible, e.g. village/ward/district) from the parasitological component will be used as a response to a vulnerability model. Once the preliminary step is finalized, the team will gather to refine the analytical model and to generate linkages between components. This exercise will be used as a platform to refine set of models and approaches proposed here.

Different approaches are proposed to be considered for analysis. Multiple analyses are considered to accommodate different context on which drivers for malaria resilience can be studied. With that, response variable(s) will be defined from one or multiple components then use data from other components as determinants or modifiers to create associations. All models will consider spatial differences of the study districts which will be defined using climate and environmental information.

The first approach will consider entomological parameters such as adult mosquito density, their infectivity (transmission) rates, resistance and information from breeding sites (number, spread and larval density) as a response. Once mapped, these will be linked to socio-demographic, socio-anthropoiogicai, parasitological and lastly HS parameters. Levels of entomological metrics are expected to be driven by rate of utilization of preventive measures, malaria infection levels, governance and institutional actions (taken by administrative team in collaboration with the HS), e.g. requesting for additional interventions such as larviciding; and lastly, human activities such as type of economic activity, choices on where to live/build a house, type of houses constructed, sources of water, knowledge and burden of disease observed in the community. This model assesses the problem from the vector aspects hence will derive actions focusing more on vector control

The second approach will take HS as the central point of deriving the understanding on the key drivers of persistence of malaria burden. Here, it is assumed that the success or failure in other components depends directly on the performance of the HS or vice versa. Taking it from here, a HS performance indicator will be built for all districts and modelled against selected/relevant indicators from all the remaining components. One of the hypothesis here could be that, if governance, institutional policies, actions and practices are not sufficiently implemented, they may result into insufficient community malaria knowledge, negative attitudes, high levels of entomological indicators, poor health seeking behaviours, poor distribution and utilization of effective interventions, and finally high burden (parasitological). This model is expected to derive perspectives that assess malaria stagnant problem from the governance actors than the beneficiaries.

The last model will approach the analysis from the perspective of availability, access and utilization of malaria preventive measures. This model will try to divide the weight of the drivers to multiple players, i.e. the HS performance, community knowledge and practices and patterns of entomological parameters for example seasonality. Since the analyses will be done using the data collected at the village level, an attempt will be made to extend the analysis to the entire district. Data from the health management information system (HMIS) which is routinely collected and reported through the Demographic Health Information system 2 (DHIS2) will be used. Such data will include test positivity rates (TPR), incidence rates, prevalence of malaria in under-fives (from MIS), school children and pregnant women using ANC data. The analyses will support development of district-wide maps of malaria burden and identify key factors to be incorporated in the ongoing efforts in the micro-stratification of malaria.

## Discussion

This protocol was used in a study that aimed at determining the reasons and factors associated with persistence of malaria in some parts of Tanzania. The regions in the north-western and southern parts of the country were selected because they showed either increasing or unchanged malaria prevalence according to the national surveys conducted from 2007 to 2016(2,32,36). The study was implemented as a cross sectional survey and covered five components which play key role in the transmission and persistence of malaria: parasitological, entomological, socio-anthropological, demographic and SES, and HS. The protocol was designed to perform a comprehensive and critical assessment of the possible drivers of resilience of malaria in these and possibly other areas with similar transmission patterns in the country.

Preliminary results show that 6,297 HHs and 28,361 individuals with median age 16yrs (IQR= 7-35yrs) were registered from the 16 villages. The population had similar structure among young individuals <20years but with more females compared to males particularly among adult individuals. Less than half (44.1%) of the HHs had family size of >4 members and >55% of the HHs owned at least one bed-net. Over 49% of individuals used bed-nets in the previous night before the survey and 43.9% of HHs had bed-nets covering two members per household. For parasitological survey, 25.8% of registered individuals (n= 7,313) were selected from 2,527 HHs (40.1%) and invited for assessment and sampling. The positivity rate (PR) by mRDTs was 33.3% ranging from 21.9% to 41.1%; while by microscopy, the PR was 20.6% and varied from 8.0% to 29.0%. Socio-anthropology interviews were conducted with a total of 1,687 heads/representatives of HHs. For qualitative surveys 32 FGDs (two from each village) and 16 IDIs (two per district) were conducted. Thirty-one health facilities were visited for the HS survey; 19.4% (n=6) were hospitals; 41.9% (n=13) health centres and 38.7% (n=12) dispensaries. In the entomological survey, 8,891 adult mosquitoes were collected, whereby *Anopheles gambiae* complex, *An. funestus* group and other mosquitoes accounted for 12.0%, 49.7% and 38.3%, respectively.

In order to successfully and timely implement study, the different components of the study were strategically planned to ensure the data collected was properly linked to prevent collection of redundant information. The entire study team was divided in small teams, which worked on their respective components in a way that allowed proper utilization data collected by the demographic and SES team. The first team (the demographic and SES group) conducted a census survey at least a week before other teams arrived in the study area. It assigned unique identification numbers to each person in the village and individuals’ data were downloaded in the electronic application that were used for data collection by other teams. This was critical to ensure that all individuals assessed in the other components had their information in the database that was used for the specific assessments done for all sub-components of the study without a need to recollect individuals’ demographics.

A comprehensive and non-traditional analysis is needed to assess the intrinsic and extrinsic drivers of the observed resilience of malaria. The analytical framework to be used includes the following, 1) developing a conceptual framework to establish the logical interlinks between the five studied components (and their attributes); 2) applying a set of models to learn the interlinks and determine associations with core drivers; and 3) proposing a set of indicators to be studied (this will be generated and refined during the process). The analysis takes into consideration the multifaceted and complex interlinks between the five components to understand these drivers while utilizing both, the quantitative and qualitative information gathered during the survey. The span of outcome studied started from the communities to the top of the ladder, which is the HS. The communities are the main beneficiaries while the actors/decision makers are within the health system. There are decision making processes within each of these two extreme ends which determine success of interventions in place; suggesting that factors behind actions of players at community level vs. health system varies. Within communities, demographic profiles and practices may be the core factors while how policies and guideline are translated into actions and innovations of players might be core factors in the HS arm. The important hypothesis that this work will use is that, there is a high variation in (almost) all core attributes even within closely located areas. The concept of “who are we dealing with here” which will be used to define the participants and develop community-based vulnerability indices is essential in fine scale assessments such as studying malaria hotspots. It is important to fully understand the type of people, their social characteristics, decision making process, their environment, assess subgroups within and between communities and tap what divides them, etc., then relate to the outcome of interest, that makes it clear to interpret associations derived from analysis.

The use of various models (with different outcome variable), set of hypothesis on how the components interacts and development of various indicators and metrics is expected to enrich the analysis, accurately estimate the effects and facilitate understanding of the contribution of each identified driver, their pathways and point out contextual factors. On the other side, the spatial analysis will feature out and quantify the variations in the realised associations and interlinks between main components or attributes within. That is an important finding to compliment stratification strategies. The analysis will be done for each district, region, zone (north-western and southern) and, lastly at country level. The cascade approach of analysis is expected to determine at which point the strength of association diminishes hence inform how future implementation should be formulated. This is deemed necessary, as some of the areas within the country are potential for malaria elimination. It is intended to test how the proposed framework is useful for this analysis, refine its components during the process of analysis (with full documentation) and share the final version (with list of indicators) to other malaria stakeholders.

The current NMCP mid-term strategic plan (MSP) 2015-2020 aims at reaching pre-elimination level by 2020. Furthermore, the strategic plan has the main goal of reducing the average country prevalence to <1%. Preliminary results from the current study have shown a high prevalence of malaria in the visited areas. This reflects that the goal of reaching the prevalence of <1% in Tanzania by 2020 and transitioning to pre-elimination may not be reached over remaining period. Various reasons may be contributing to that situation, among them being wide transmission diversity which may have an impact on the performance of NMCP in attaining its MSP goals. In response to that, during their midterm review of the MSP, NMCP introduced supplementary plan with an objective of facilitating achievement of MSP goal by 2020. Among the revised strategies, is stratification and deploying intervention based on the local burden of malaria and making surveillance, monitoring and evaluation a core intervention as recommended by WHO (54). The overall aim is to enable NMCP to monitor progress in malaria control and produce country specific evidence-based stratification. The findings of the proposed analysis will be key in developing a framework that will be routinely used by NMCP to continuously re-stratify malaria burden using data to be generated from intensified surveillance.

Analysis and results from this study will determine factors which might be potentially responsible for persistence of malaria and hotspots in the study areas and possibly other parts of Tanzania. The proposed triangulated analysis of the five components, rather than the traditional methodology of focusing on one metric, is expected to highlight potential drivers, their complex interactions and map spatial heterogeneity which will be an eye-opener to better understanding of determinants of malaria hotspots. This study is expected to provide an important framework and data which will guide future studies and malaria surveillance in Tanzania and other malaria endemic countries. The findings of the study will inform the NMCP and its stakeholders on stratified and malaria targeted control interventions required to push malaria further down to reach pre-elimination target by 2020 and elimination by 2030. The protocol can be modified and customized, and will be useful to other countries which might be interested in undertaking similar assessments.

## Data Availability

The datasets which will be analysed for the current study are available at the NMCP, and can be shared through the corresponding author on reasonable request.

**List of Abbreviations**
ADDO: Accredited drug dispensing outlet
ANC: Antenatal clinic
CDC: Centres of Diseases Control and Prevention of US
DBS: Dried blood spot
DED: District executive director
*dhfr*: Dihydrofolate reductase gene
DHIS2: Demographic health information system 2
*dhps*: Dihypteorate synthase gene
DMFP: District Malaria Focal Person
DMO: District medical officer
DNA: Deoxyribonucleic acid
ELISA: Enzyme-linked immunosorbent assay
FDG: Focus group discussion
G6PD: Glucose 6-phosphate dehydrogenase
gDNA: Genomic deoxyribonucleic acid
GPS: Global positioning system
HHs: House holds
HMIS: Health management information system
HRP2: Histidine rich protein 2
HS: Health system
IPD: inpatient department
IPTp: Intermittent presumptive treatment in pregnancy
IQR: interquartile range
IRS: Indoor residual spraying
ITNs: Insecticide treated nets
ITS: internal transcribed spacer
KAP&B: Knowledge, attitude, practice and beliefs
*kdr*: knock down resistance gene
LLINs: Long-lasting insecticidal nets
*mdn*: multidrug resistance 1 gene
MIP: Molecular inversion probes
MIS: Malaria indicator survey
MRCC: Medical research coordinating committee
mRDT: Malaria rapid diagnostic test
MSP: mid-term strategic plan
NMCP: National malaria control programme
ODK: Open data kit
OPD: Outpatient department
PCR: Polymerase chain reaction
PET: Photo-induced electron transfer
*Pf*: *Plasmodium falciparum*
*Pf-*CSP: *Plasmodium falciparum* circumsporozoite protein
PO-RALG: President’s office, regional administration and local government authority
PR: Positivity rate
RCH: Reproductive and child health
rDNA: Recombinant deoxyribonucleic acid
SES: Socio-economic status
SMPS: School malaria prevalence survey
SNPs: Single nucleotide polymorphisms
SP: Sulphadoxine/pyrimethamine
SPAM: Service provision assessment for malaria
SSA: sub Saharan Africa
TDHS: Tanzania demographic and health survey
THMIS: Tanzania HIV and malaria indicator survey
TPR: Test positivity rate
WBC: White blood count
WHO: World health organization

## Declarations

### Ethical approval and consent to participate

This study obtained ethical approval from the Medical Research Coordinating Committee (MRCC) of the National Institute for Medical Research (NIMR/HQ/R.8a/Vol.IX/2551). Furthermore, permission to conduct the study in the selected regions was sought from the President’s Office, Regional Administration and Local Government Authority (PO-RALG) and Regional, District and village authorities. Informed consent/assent was sought before conducting the demographic survey or including the participants into the different components of the study as well as before inserting mosquito traps into the houses.

### Consent for publication

Not applicable

### Authors’ contribution

DSI, MGC, CIM, SFR, FC, FF, BPM, MDS, LN, ISM, EL, YAD, BMB, RAM and FM made substantial contributions to the conception and design of the protocol; DSI, MGC, SFR, FC, FF, MPM and FM designed the analysis and data interpretation plan. DSI, MGC, CIM, SFR, FC, FF, BPM, MDS, DPC, LN, ISM, EL, WHM, ADM, VB, MDS, YAD, BMB, PMH, RAM, MCM, RM, AM, FM, RN and AM contributed in drafting and revising the protocol. All authors reviewed and approved the final version of the manuscript.

## Acknowledgments

The study team would like to thank all study participants for their contribution in the study. Special appreciation goes to the President’s Office Local Government and Regional Authorities and the village, district and region authorities; particularly village Leaders, District Executive Directors (DEDs), District medical Officers (DMOs) and District Malaria Focal Persons (DMFPs) for supporting the team during the entire study period.

The team would also like to acknowledge the contribution of the research scientists and assistants; Gasper Lugela, Francis Chambo, Tilaus Gustav, Juma Akida, Mwanaidi Mtui, Caroline Minja, Maimuna Ahmed and Richard Makono from the Parasitology team. The demographic team included; Gineson Nkya, Richard Malisa, Raphael Charles, Respigi Kiwango, Evord Kimario, Lifoba Mosoud, Stumai George, Ali Idris, Ibrahim Materego, Neema Barua, and Michael Makange. Cloud Tesha, Emmanuel Chagoha and Happyness Jeremiah were members of the socio-anthropology team. The entomology team included Martin Zuakuu, Adam Mgaya, Hashim Ally, Heri Bakari, Alex John, Neema Richard, Rukia Ahmed and Israel Msangi. The technical, administrative and logistic support rendered by the Finance department at Tanga Centre (Lydia Lugomola, Derick Maira, Joseph Said and Selemani Mandia) and NMCP (Abdallah Kajuna and Judith Kirama); drivers (Seth Nguhu, Francis Mkongo, Saidi Maivaji, Silvester Msamila, Eliwasiri Mmbaga, Thomas Semdoe, Athumani Simba and Ally Mshana) and others (Robert Mhilu, Salome Ngoda, Elfrida Mosha, Haruna Mrisho, Beatrice Semng’indo and Rehema Mtibusa) is highly appreciated.

We are grateful to WHO through Tanzania Country Office for providing technical support. The National Institute for Medical Research is thanked for the support and providing permission to publish this protocol.

